# Real-time detection of COVID-19 epicenters within the United States using a network of smart thermometers

**DOI:** 10.1101/2020.04.06.20039909

**Authors:** S.D. Chamberlain, I. Singh, C. Ariza, A. Daitch, P. Philips, B. D. Dalziel

## Abstract

Containing outbreaks of infectious disease requires rapid identification of transmission hotspots, as the COVID-19 pandemic demonstrates. Focusing limited public health resources on transmission hotspots can contain spread, thus reducing morbidity and mortality, but rapid data on community-level disease dynamics is often unavailable. Here, we demonstrate an approach to identify anomalously elevated levels of influenza-like illness (ILI) in real-time, at the scale of US counties. Leveraging data from a geospatial network of thermometers encompassing more than one million users across the US, we identify anomalies by generating accurate, county-specific forecasts of seasonal ILI from a point prior to a potential outbreak and comparing real-time data to these expectations. Anomalies are strongly correlated with COVID-19 case counts and may provide an early-warning system to locate outbreak epicenters.

**One Sentence Summary:** Distributed networks of smart thermometers track COVID-19 transmission epicenters in real-time.

## Main Text

Epidemic dynamics of the coronavirus disease (COVID-19) have varied widely across the globe, since the outbreak emerged from Wuhan, China in December 2019 (*1*). Countries with proactive management and widespread testing have more effectively limited spread (i.e. ‘flattening the curve’), whereas countries with limited response, such as the United States, have experienced steeper growth rates in cases (*2*). Syndromic monitoring systems—collecting information about symptoms often prior to, or instead of, interaction with the formal healthcare system—have played a pivotal role in successfully mitigating COVID-19 spread in countries such as Taiwan and South Korea (*3*). Rapid syndromic detection also played a role in curtailing the 2014-2016 Ebola Outbreak in West Africa (*4*), where speed of detection was important in reducing outbreak intensity (*5*).

Networks of geolocated, user-generated physiological measurements hold the potential for improved tracking and prediction of outbreak epicenters (*6, 7*). Data from these networks are typically less specific, but more sensitive, than formal surveillance, and are often available more rapidly, because formal surveillance is constrained by testing speed (*5*) and/or time for record aggregation (*8*). Data from syndromic monitoring networks can be physiologically grounded in disease processes (e.g. fever), in contrast with internet-based proxies, such as search engine logs, that are often used as a substitute for real-time syndromic data (*9*). Physiological data from connected devices, such as thermometer data, or elevated heart-rate data (*10*), are thus a potentially valuable resource for outbreak response, and, at present, may be able to detect COVID-19 symptoms in the absence of widespread and rapid testing in the US.

Here, we outline a method to identify illness incidence anomalies using a geospatial network of smart thermometers, where county-scale anomalies are flagged in real-time. We then use the method to identify COVID-19 outbreaks in the continental United States. Our anomaly detection method follows three core steps: 1) Generate county-specific forecasts of influenza-like illness (ILI) from a time point prior to a potential outbreak, 2) compare real-time thermometer-derived ILI to forecast expectations when new data is aggregated daily, and 3) flag anomalous ILI values by evaluating the probability that the current signal is driven by regular seasonal influenza. In this case, we flag ILI values that exceed the 97.5% percentile of expected influenza trajectories.

We track real-time ILI and create ILI forecasts using data collected from a network of smartphone-connected personal thermometers managed by Kinsa, Inc. We track ILI in the network by classifying users with elevated temperature readings over multiple days as having influenza-like illness (see Methods). This sensor network records the temperature and approximate geo-location when a user takes their temperature (e.g. during an illness episode). These recordings store temperature readings, via Kinsa’s smartphone app, and locations are logged via GPS location or IP address. Readings are aggregated to county-scale ILI and anonymized. The temperature readings are used to construct an ILI signal that is highly correlated to Center for Disease Control and Prevention (CDC) ILI nationally (*r* > 0.95) and across CDC regions (*r* range 0.70-0.94), and these signals have been demonstrated to improve regional ILI surveillance and forecasting (*6, 7*). We construct the ILI signal at the county-scale, allowing identification of anomalous ILI incidence at the scale of individual cities. More information on constructing illness incidence from the thermometer network can be found in the Supplemental Methods.

Our forecast builds upon the findings that individual cities have predictable, distinct epidemic intensity patterns governed by both population size and climate (*11, 12*). As population size and climate change relatively slowly from year-to-year, this allows the use of the county-specific, daily network data to learn the typical seasonal flu transmission patterns of a region. From this we construct local ILI forecasts that form the basis for real-time anomaly detection. Following (*13*), we estimate a county’s daily reproductive number (*R*_*t*_), the average number of secondary cases that each infected individual would infect if the conditions remained as they were, at time *t* by solving the following equation:

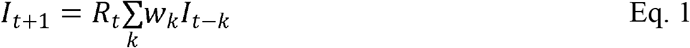

where *I*_*t*_ is Kinsa-derived ILI incidence, and *w*_*k*_ is the infectivity profile for influenza, representing the probability that an individual who becomes infected on day *t* acquired the infection from an individual who became infected on day *t-k*. We approximate *w*_*k*_ by a gamma distribution with a mean of 2.5 and variance of 0.7 days (*14*). The summation term in equation (1) thus represents “effective incidence” assumed to be experienced by a susceptible individual on day *t*. For each county, we estimate the median *R*_*t*_ for each day-of-year by first calculating daily *R*_*t*_ for back to August 1, 2016. We then forecast ILI forward from March 1^st^, 2020, by using the median *R*_*t*_ corresponding to day *t*, to estimate *I*_*t+1*_ and forward propagate these predictions 12-weeks (84 days) ahead. We generate an ensemble of 100 influenza predictions by inserting Gaussian noise into the starting values of effective incidence at the point of prediction. The scale of noise varies across regions, determined from the residual ILI after detrending the past time series using a two-week centered, rolling mean. See Methods for more detail on the nation-wide and region-specific implementations of this approach.

We compare the real-time ILI to these influenza trajectories to identify localized anomalies, daily. We flag anomalous ILI when the daily, real-time data exceeds the 97.5% percentile of the expected influenza forecast ensemble. We define *anomaly incidence* as the difference between the real-time thermometer ILI and the 97.5% percentile drawn from the influenza forecast ensemble. To validate this method, we apply our approach to detect COVID-19 anomalies by forecasting expected influenza starting on March 1^st^, 2020, before widespread outbreaks were underway (*2*). We assess detection quality by comparing *anomaly fever counts* to state- and county-level COVID-19 confirmed cases aggregated from a variety of government sources (*15*). Anomaly fever counts are calculated from cumulative anomaly incidence multiplied by the estimated user base of a region.

The skill of the ILI forecast in the anomaly detection system compares favorably with the Carnegie Mellon (CMU) Epicast and Stat models (*16*) at CDC region and national scales (Fig. 1A). To calculate these forecast error rates, we compare CMU forecasts to CDC ILI, while comparing anomaly detection forecasts to thermometer-based ILI, given that these models are trained on different data sources. In contrast to other ILI forecasts, our models do not need to be initialized with now-cast predictions, (i.e. inferred initial conditions) as thermometer-based ILI, upon which the model is trained, is available in real-time. Additionally, our forecast errors do not appear to increase much past the 5-week forecast horizon, suggesting the ILI forecasts in the anomaly detection system are stable at long-time horizons (Fig. 1A). Forecast error rates display seasonality, where forecasting errors are higher in winter months during periods of increased illness incidence, and lower in the spring and summer (Fig. S1).

**Figure 1:**
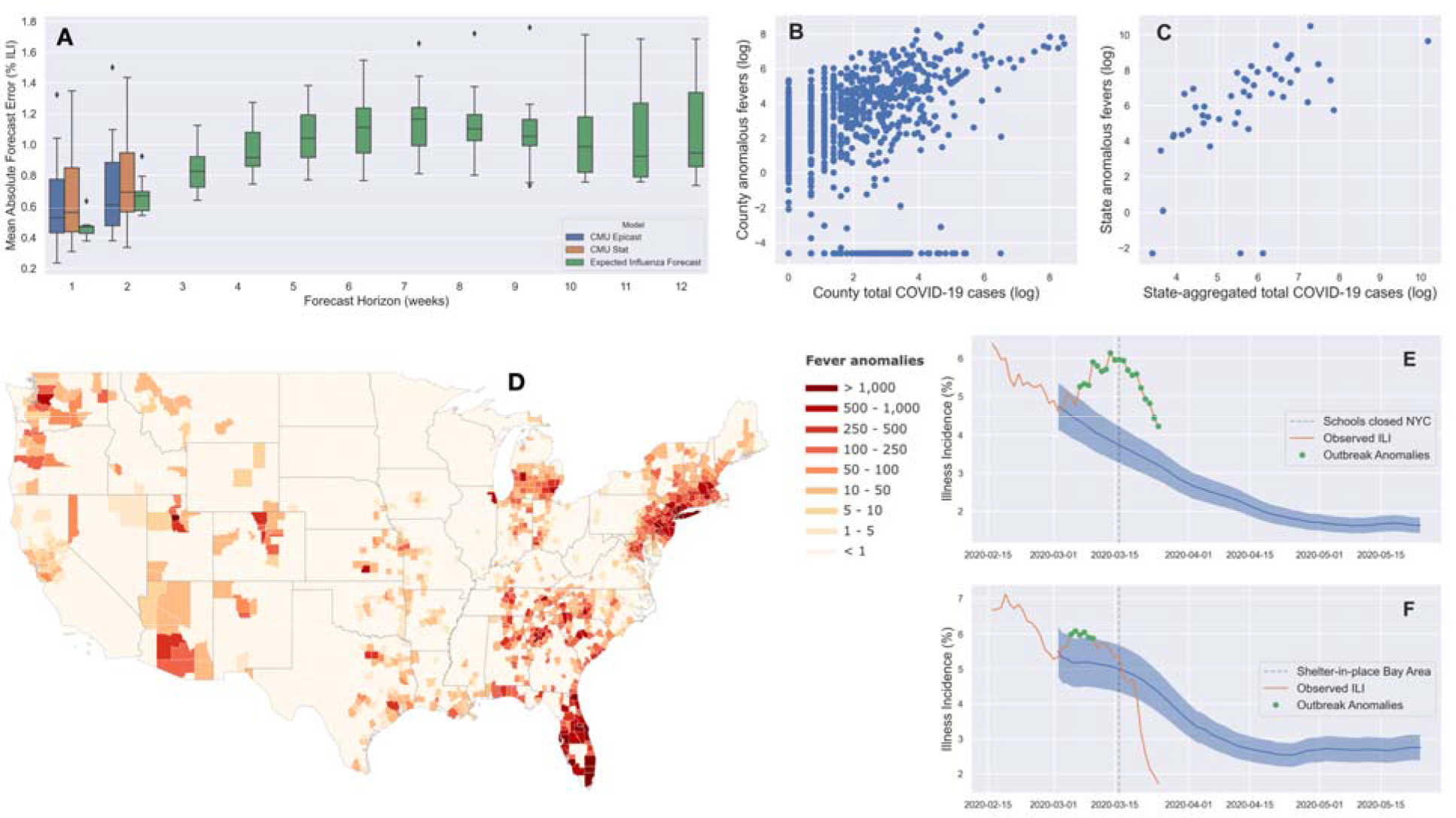
Model comparisons (**A**) at CDC region and national scales between the expected influenza and Carnegie Mellon forecasts. Correlations between detected fever anomalies and total confirmed COVID-19 cases at the (**B**) county- and (**C**) state-scale. These comparisons were made across 1321 counties with at least one confirmed case. (**D**) Map of anomalous fever estimates on March 24^th^, 2020, where anomalous fevers are estimated from cumulative anomaly incidence multiplied by the county user base. Anomaly incidence detections in (**E**) Brooklyn, NY, USA and (**F**) Santa Clara, CA, USA where school closures and shelter-in-place orders were implemented. Here, the blue line and shaded area are the median expected influenza forecast and 2.5-97.5^th^ percentile of the forecast ensemble, respectively.

Our detected anomalies correlate strongly to positive COVID-19 cases at both county (Fig. 1B) and state scales (Fig. 1C), validating this method for the rapid detection of COVID-related illness anomalies. Specifically, we find significant correlations between total confirmed COVID-19 cases and total fever anomalies at the county (Fig. 1B: *r* = 0.54, *P* < 0.0001) and state scales (Fig. 1C: *r* = 0.55, *P* < 0.0001). We restrict this analysis to counties with at least one confirmed COVID-19 case, given that a lack of confirmed cases could be due to either true COVID-19 patterns or an absence of testing and reporting. We do not observe anomalous fevers in three states where COVID-19 cases are confirmed; Minnesota, Wisconsin, and South Dakota. County-level total anomalous fevers also correspond to the spatial distribution of major known outbreak centers, with hotspots in the Seattle Area, San Francisco Bay Area, New York Metro Area, and Florida (Fig. 1D).

We provide an example for how this method is applied in real-time for Brooklyn, NY, USA (Fig. 1E). Here, we forecast expected influenza trends from March 1^st^, 2020 before large-scale outbreaks were occurring in the New York Metro Area (*2*), and we observe a strong divergence from expected influenza trajectories (Fig. 1E). However, we also observe an inflection back toward declining ILI shortly after social distancing efforts were enacted (Fig. 1E), beginning with school closures on March 16^th^ and a later ‘stay-at-home’ order on March 20^th^, 2020. We observe even sharper declines in ILI post-social distancing in Santa Clara, CA, where a ‘shelter-in-place’ order was enacted on March 16^th^, 2020 (Fig. 1F).

Our estimates of anomaly incidence are likely impacted by social distancing in these cases, as distancing should also reduce influenza transmission. We are thus likely underestimating anomaly incidence after social distancing is enacted, given that our model assumes typical seasonal influenza transmission patterns. We therefore explore the sensitivity of our method by reducing *R*_*t*_ values used in the influenza forecast for all days after social distancing efforts were implemented. In Brooklyn NY (Fig. 2A), dropping future *R*_*t*_ values by 25% reduces the trajectory of seasonal influenza, leading to an increased magnitude of anomaly incidence (Fig. 2A). In Santa Clara County, without accounting for social distancing we observe only a short period of detected anomalies, where real-time ILI falls below the expected range of seasonal influenza (Fig. 1F). Here, reducing *R*_*t*_ following the ‘shelter-in-place’ order leads to additional anomaly detections post-social distancing (Fig. 2B). Similarly, following school closures in Miami-Dade County, real-time ILI values fell below anomalous levels by late March (Fig. S2), though accounting for reduced influenza transmission leads to more anomaly detections (Fig. 2C). In these case examples, we assumed *R* reduced by 25% given social distancing directives. While the true value cannot be estimated at present, it likely varies with both policy implementation and adherence. These findings suggest social distancing plays an important role in the quality of anomaly detection, and future research should address the impact of social distancing on influenza transmission rates.

**Figure 2:**
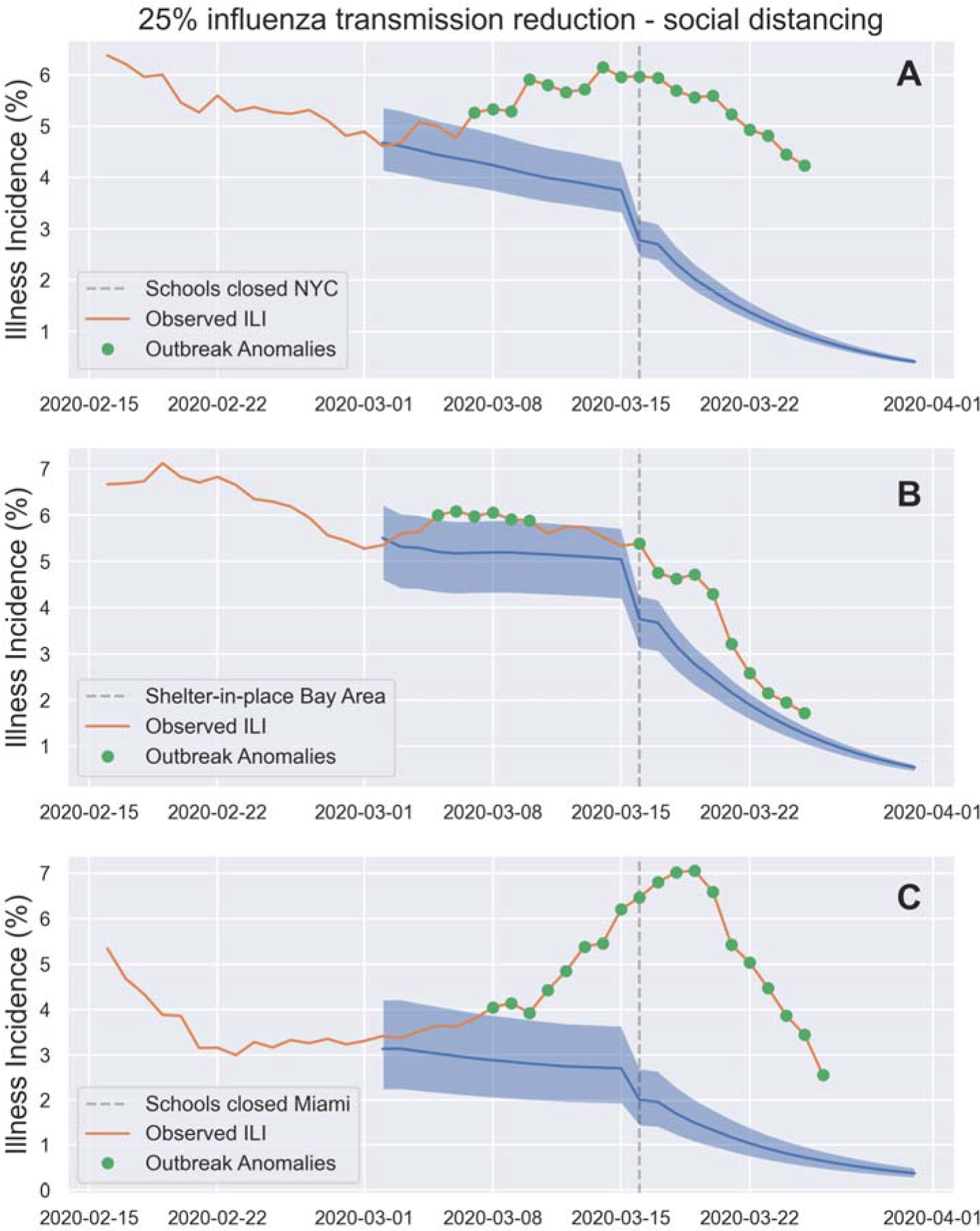
Influence of social distancing-based reductions of influenza transmissivity (*R*_*t*_) on anomaly detection in (**A**) Brooklyn, New York, (**B**) Santa Clara, California, and (**C**) Miami, Florida. Reductions of influenza *R*_*t*_ by 25% cause additional anomaly detections in Santa Clara and Miami and increase the magnitude anomaly incidence detections in Brooklyn.

Working in synergy with the formal healthcare system, high-throughput signals from distributed syndromic monitoring networks, such as we describe here, could play a decisive role in managing local outbreaks, and alter the trajectories of pandemics. These networks may be of particular use for situations where rapid testing and isolation are key to pandemic control (*3, 4*), such as the current COVID-19 outbreak, where the absence of widespread and rapid testing have led to a massive growth in cases worldwide.

## Data Availability

The anomaly detection and forecasting code can be accessed on GitHub, and all examples here are reproducible (https://github.com/kinsahealth/therm_anomaly_detection).

https://healthweather.us/

## Acknowledgements

The anomaly detection and forecasting code can be accessed on GitHub, and all examples here are reproducible (https://github.com/kinsahealth/therm_anomaly_detection). We thank C. Jessica Metcalf for her input on an initial draft of this manuscript.

## Author Contributions

SDC conceived and implemented the anomaly detection approach with feedback from BDD and PP. IS conceived of and designed the Kinsa products, including its use as syndromic early warning and monitoring systems. PP, CA, AD, and SDC designed and managed the real-time illness signal and geospatial platform. SDC wrote the manuscript with feedback from all authors.

## Ethics Statement

This work is not a clinical trial and is instead a population-level observational study, where all user information is anonymized. More specifically, this study uses aggregated temperature readings to estimate the prevalence of influenza-like illness at the scale of United States counties. Raw measures are temperature, time of reading, and approximate geolocation from smart thermometer readings, where user information is anonymized and aggregated to whole population influenza-like illness at the scale of US counties. It therefore does not use individual, private or personally identifiable information. Given the nature of this data, it does not require IRB review.

## References and Notes

1. “Coronavirus Disease 2019 (COVID-19) – Situation Report – 51” World Health Organization (2020; https://www.who.int/docs/default-source/coronaviruse/situation-reports/20200311-sitrep-51-covid-19.pdf?sfvrsn=1ba62e57_10)

2. Johns Hopkins University, Coronavirus Resource Center (2020; https://coronavirus.jhu.edu/map.html; data available at https://github.com/CSSEGISandData/COVID-19)

3. C. J. Wang, C. Y. Ng, R. H. Brook, Response to COVID-19 in Taiwan: Big data analytics, new technology, and proactive testing. JAMA (2020) doi:10.1001/jama.2020.3151

4. L. A. McNamara et al., “Ebola surveillance – Guinea, Liberia, and Sierra Leone” MMWR supplements 65 (2016; https://www.cdc.gov/mmwr/volumes/65/su/su6503a6.htm)

5. P. Nouvellet, T. Garske, H. L. Mills, G. Nedjati-Gilani, W. Hinsley, I. M. Blake, M. D. Van Kerkhove, A. Cori, I. Dorigatti, T. Jombart, S. Riley, The role of rapid diagnositics in managing Ebola epidemics. Nature 528, 7580 (2015).

6. A. C. Miller, I. Singh, E. Koehler, P. M. Polgreen, A smartphone-driven thermometer application for real-time population-and individual-level influenza surveillance. Clinical Infectious Diseases 67, 388–397 (2018).

7. A. C. Miller, R. A. Peterson, I. Singh, S. Pilewski, P. M. Polgreen, “Improving state-level influenza surveillance by incorporating real-time smartphone-connected thermometer readings across different geographic domains” in Open Forum Infectious Diseases (Oxford University Press, 2019).

8. Center for Disease Control and Prevention, Weekly U.S. Influenza Surveillance Report (2020; https://www.cdc.gov/flu/weekly/index.htm)

9. S. Yang, M. Santillana, S. C. Kou, Accurate estimation of influenza epidemics using Google search data and ARGO. Proceedings of the National Academy of Sciences 112, 14473–14478.

10. J. M. Radin, N. E. Wineinger, E. J. Topol, S. R. Steinhubl, Harnessing wearable device data to improve state-level real-time surveillance of influenza-like illness in the USA: a population-based study. The Lancet Digital Health

11. B. D. Dalziel, S. Kissler, J. R. Gog, C. Viboud, O. N. Bjornstad, C. J. E. Metcalf, B. T. Grenfell, Urbanization and humidity shape the intensity of influenza epidemics in US cities. Science 6410, 75–79 (2018).

12. B. D. Dalziel, B. Pourbohloul, S. P. Ellner, Human mobility patterns predict divergent epidemic dynamics among cities. Proceedings of the Royal Society B: Biological Sciences 280, 1766 (2013).

13. A. Cori, A new framework and software to estimate time-varying reproductive numbers during epidemics. American Journal of Epidemiology 178, 1505–1512 (2013).

14. F. Carrat, E. Vergu, N. M. Ferguson, M. Lemaitre, S. Cauchemez, S. Leach, A. J. Valleron, Time lines of infection and disease in human influenza: A review of volunteer challenge studies. American Journal of Epidemiology 167, 775–785 (2008).

15. L. Davis, Corona Data Scrapper (2020; https://github.com/lazd/coronadatascraper)

16. Carnegie Mellon University, DELPHI Group (2020; https://github.com/cmu-delphi/delphi-epidata)

